# Anxiety-impulsivity subtypes in adolescent internalizing disorder are characterized by distinguishable neurodevelopmental, neurocognitive and clinical trajectory signatures

**DOI:** 10.1101/2021.10.30.21265692

**Authors:** Huaxin Fan, Nanyu Kuang, Xinran Wu, Gechang Yu, Tianye Jia, Barbara J. Sahakian, Trevor W. Robbins, Gunter Schumann, Jianfeng Feng, Benjamin Becker, Jie Zhang

## Abstract

**Background:** Anxiety and impulsivity represent transdiagnostic pathology dimensions yet their interaction and contribution to emotional disorders in adolescence and to disease development remain controversial, and previous studies indicate heterogeneity within the broad category of internalizing disorders.

**Methods:** A combination of hierarchical and non-hierarchical clustering strategies was employed to determine impulsivity-related subtypes (based on the facets of negative urgency, lack of planning, lack of perseverance, sensation seeking and positive urgency in UPPS-P scales) in a large cohort of adolescents with internalizing disorders (n=2437) from Adolescent Brain Cognitive Development (ABCD) Study. Linear mixed-effect models were employed to determine cortical thickness alterations of the subtypes.

**Results:** Data-driven clustering identified two distinct subtypes of internalizing patients (subtype 1/subtype 2) with comparable levels of increased anxiety yet distinguishable levels of impulsivity, i.e., enhanced (subtype 1) or decreased (subtype 2) compared to healthy controls. Subtype 1 was further characterized by thicker prefrontal and temporal cortical regions involved in regulatory control and fear processing, while subtype 2 did not demonstrate significant thickness alterations. The differential neuroanatomical profiles remained stable over the two-year follow-up, while the two subtypes had different neurodevelopmental trajectories. Subtype 1 additionally reported more psychopathology and dysfunctionality including higher suicidal ideation, depressive symptoms and transition rates to externalizing disorders during follow-up as well as impaired neurocognitive and educational performance compared to subtype 2. Moreover, for subtype 1, anxiety at baseline (9-10 years) was significantly positively associated with impulsivity (lack of perseverance) at 2-year follow-up, while in subtype 2, baseline anxiety was significantly negatively associated with impulsivity (sensation seeking) at 2-year follow-up.

**Conclusions:** Our results demonstrate an impulsivity-dependent heterogeneity in adolescent internalizing disorders, with high-impulsivity patients being characterized by neurodevelopmental delay at the neural and cognitive levels. Individuals with elevated impulsivity are at a greater risk to develop behavioral dysregulation over the following two years and may thus require specific early interventions.

## Introduction

The conceptualization of psychiatric disorders as distinct diagnostic entities has been challenged by high comorbidity rates on the symptomatic level and increasing evidence for broad transdiagnostic meta-dimensions of psychopathology, such as internalizing and externalizing or impulsivity and compulsivity (Caspi et al 2020; Pasion and Barbosa 2019; Kotov et al 2017; Lees et al 2021; Robbins et al 2012; Romer et al 2021). This conceptualization offers advantages in terms of determining common etiological processes and neurobiological dysregulations (American Psychiatric Association 2013; Cuthbert and Insel 2013). Within the context of this dimensional conceptualization disorders are regarded as extreme points on continua that span a range of emotional and behavioral functions (Krueger 1999) [see e.g. also the Research Domain Criteria (RDoC) (Cuthbert and Insel 2013)] rather than traditional discrete diagnostic entities (Hudziak et al 2007).

Emotional or internalizing disorders represent the largest group of psychiatric disorders - encompassing primarily affective and mood disorders - which are characterized by exaggerated levels of anxious distress. In addition to anxiety, impulsivity - the tendency to act inappropriately and prematurely despite adverse outcomes also represents a major dimension of psychopathology (Cosi et al 2011; Cummings et al 2014; Whiteside and Lynam 2001). Whereas both dimensions have been increasingly associated with robust neurobiological dysregulations across diagnoses (Xu et al 2020; Linke et al 2021;

Newman et al 2016; Owens et al 2020), anxiety and impulsivity have usually been examined separately in internalizing or externalizing disorders, respectively. Accumulating evidence suggests that impulsivity may play a role in internalizing disorders, while findings on the relationship between anxiety and impulsivity in internalizing disorders remain inconsistent (Jakuszkowiak-Wojten et al 2015) such that the traditionally reported inverse association (Apter et al 1993; Taylor et al 2008) has been challenged by an accumulation number of recent studies reporting a positive relationship (Cosi et al 2011; Favaloroa and Moustafa 2020; Moustafa et al 2017; Yu et al 2020).

While exaggerated anxiety and impulsivity have been identified as key diagnostic dimensions in adult samples, most psychiatric conditions, including internalizing disorders, emerge already during adolescence (Castellanos-Ryan et al 2016; Rakesh et al 2020), a period characterized by fundamental processes of brain development and maturation (Shaw et al 2008), particularly synaptic pruning and myelination (Merz et al 2018). The neuromaturational changes are mirrored in the macroscale architecture of the brain particularly cortical thickness, which increases during childhood, decreases during adolescence and stabilizes during early adulthood (Huttenlocher 1979; Natu et al 2019; Shaw et al 2008). Dysregulation of neuromaturation of cortical thickness has been closely linked with the development of psychiatric disorders (Whittle et al 2020) and impaired cognitive performance (Shaw et al 2006). The developmental trajectories of brain systems differ, such that the “dual systems” or “maturational mismatch” theories hypothesize that regulatory prefrontal cortical regions undergo prolonged maturational changes while subcortical regions involved in reward and emotion processing mature earlier, leading to a developmental mismatch which in turn promotes emotion regulation and behavioral problems during adolescence (Casey et al 2008; Powers and Casey 2015; Steinberg 2008). Therefore, examining the interactions between anxiety and impulsivity and associations with adolescent neuromaturational trajectories may allow the determination of broad subtypes of neuropathological dysregulations at the core of psychiatric disorders in later life.

We aimed at determining the relationship between anxiety and impulsivity in a large sample of early adolescents with internalizing disorders. Specifically, we first clustered adolescents with pure internalizing disorders according to their levels of impulsivity using a data-driven approach. Next, we examined the differences between the groups on the neurobiological level by determining distinct brain morphological profiles in the subtypes. Subsequently, we explored the relationship between the dimensions of anxiety and impulsivity within the identified subtypes. Finally, we examined the differences of neurocognitive and educational performance between groups and differences between the subtypes at the clinical predictive level in terms of pathological trajectories during follow-up. Given that anxiety and impulsivity are major dimensions for the development of adolescent psychopathology (Cosi et al 2011) and impulsivity has been linked with brain morphological neurodevelopmental dysregulation (Owens et al 2020; Newman et al 2016; Zhu et al 2020) we hypothesized that the interactions at the symptomatic level should be reflected in terms of cortical thickness, specifically impulsivity-associated dysregulations in prefrontal regions, and that these alterations remain stable or increase over a development trajectory spanning the age between 10 to 12. Given the clinical challenge of predicting behavioral dysregulation in adolescence and previous findings suggesting that impulsive-anxious adolescents show a higher inclination to suicidality and depression (Askenazy et al 2003), we specifically focused on the development of suicidality, depression and behavioral dysregulation in terms of externalizing symptomatology.

## Methods and Materials

### Participants

A sample of 11878 children aged 9-10 years was obtained from Data Release 3.0 of the Adolescent Brain Cognitive Development (ABCD) study (https://abcdstudy.org/scientists/data-sharing/). The ABCD study is a large longitudinal study that recruited children from 21 research sites across the USA (Casey et al 2018), and currently incorporates clinical, behavioral, cognitive, and multimodal neuroimaging data from baseline, 1-year and 2-year follow-up assessment (https://abcdstudy.org/scientists/protocols/). The ABCD study group obtained written and oral informed consent from parents and children, respectively (Auchter et al 2018). Details on the protocols and assessments are provided in corresponding publications from the ABCD study group (Barch et al 2018; Hagler Jr et al 2019; Casey et al 2018).

For the present study we excluded participants with missing data of corresponding measures or covariates and with bipolar disorder, disruptive mood dysregulation disorder, psychotic disorder, alcohol use or substance use disorder for the internalizing patients, as well as data which did not pass quality control for neuroimaging data.

### Measures

#### Categorical psychiatric diagnosis

Categorical psychiatric diagnoses were obtained via parent report using the structured computerized Kiddie Schedule for Affective Disorders and Schizophrenia for DSM-5 (KSADS-5) (Barch et al 2018). For the present analysis lifetime diagnoses for the respective specific or unspecific disorders were employed. Based on the definitions of transdiagnostic dimensions of psychopathology in recent studies (Romer et al 2021; Caspi et al 2020; Lees et al 2021; Pasion and Barbosa 2019), two broad diagnostic families in our analyses were determined as internalizing disorders (depressive disorder, agoraphobia, panic disorder, specific phobia, separation anxiety disorder, social anxiety disorder, generalized anxiety disorder and post-traumatic stress disorder) and externalizing disorders (attention-deficit/hyperactivity disorder, oppositional defiant disorder and conduct disorder).

Our initial analyses focused on individuals with pure internalizing disorders while excluding subjects with comorbid externalizing disorders to control for the potential influence of comorbid externalizing conditions (Figure 1A). Individuals classified into this group were required to (1) fulfill the criteria for at least one internalizing disorders, while (2) comorbid externalizing disorders, bipolar disorder, disruptive mood dysregulation disorder, psychotic disorder, alcohol use disorder or substance use disorder led to exclusion. To test the robustness of our findings we repeated the analyses in all internalizing disorders patients (including those with comorbid externalizing conditions) (Figure 1A). Individuals without any lifetime diagnosis were classified as healthy controls (HC) and served as a reference group. Numbers of subjects and section procedures are shown in Figure 1B.

**Figure 1.**
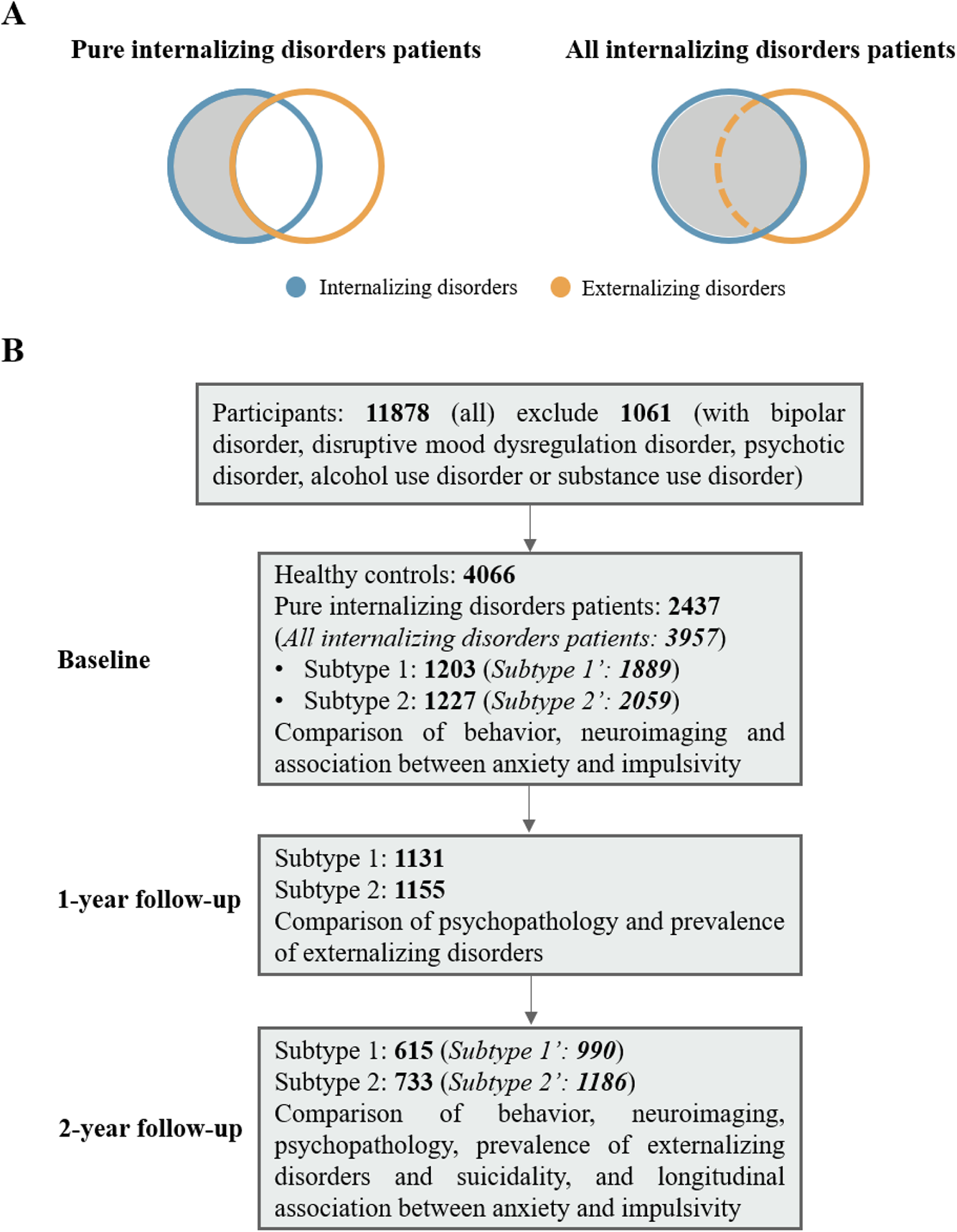
Selection of subjects. (A) Visualization of selection of pure internalizing disorders patients and all internalizing disorders patients. (B) Flowchart showing selection of subjects and number of participants in each subgroup analysis. Our initial analyses focused on individuals with pure internalizing disorders while excluding subjects with comorbid externalizing disorders to control for the potential influence of comorbid externalizing conditions. To test the robustness of our findings we repeated the analyses in all internalizing disorders patients (italic in brackets) including those with comorbid externalizing conditions.

#### Trait impulsivity and sub-facets

Trait impulsivity and its sub-facets were assessed by means of the validated UPPS model of impulsivity (UPPS-P Impulsive Behavior Scale, Barch et al 2018) encompassing the sub-facets Negative and Positive Urgency (nu and pu, behavioral dysregulations in the context of negative or positive emotions, respectively), Lack of Premeditation and Perseverance (lopl and lope, indexed by tendencies to act without planning or quit difficult tasks, respectively), and Sensation Seeking (ss, seeking arousing and stimulating activities).

#### Anxiety, depression and behavioral problems scores

Levels of anxiety were derived from the Child Behavior Checklist (CBCL) a widely used parent-report of child and adolescent behavior (Barch et al 2018) by capitalizing on raw scores of the Anxiety Problems scale which is aligned with DSM-anxiety symptoms. We additionally included the Depressive Problems as an index of depressive symptom load and other subscales (Externalizing Problems, Rule-Breaking Behavior, Aggressive Behavior, ADHD Problems, Oppositional Defiant Problems, Conduct Problems) to represent children’s behavioral problems.

#### Behavioral inhibition and activation

Gray’s Reinforcement Sensitivity Theory (Gray 1987) proposes two neurobiological rooted motivational systems, the Behavioral Inhibition System (BIS) which determines responses to punishment and generates anxious emotions, and the Behavioral Activation System (BAS) which determines sensitivity to rewards and has been related to impulsivity (Gray 1987). Four sub-facets are included in the validated BIS/BAS scale, three for behavioral activation: Drive (basdr, intensity of goal directed behavior), Fun seeking (basfs, enjoyment for its own sake, spontaneity), and Reward Responsiveness (basrr, excitement over reinforcing outcomes) and one for behavioral inhibition (bis, e.g., worry, fearfulness) (Barch et al 2018).

#### Neurocognitive functioning

Neurocognitive functioning in the domains of language vocabulary knowledge, attention, cognitive control, executive function, episodic memory, working memory, processing speed and flexible thinking as well as composite scores for crystallized intelligence, fluid intelligence and total intelligence (Weintraub et al 2013) were derived from the validated NIH toolbox.

#### Structural image acquisition and preprocessing

T1-weighted structural MRI (sMRI) data were collected on 3T MRI systems (Siemens Prisma, General Electric MR 750, Philips). Detailed information of acquisition is described in (Casey et al 2018). SMRI data preprocessing was completed by the ABCD study according to standardized processing pipelines (Hagler Jr et al 2019). Cortical surface reconstruction and subcortical segmentation were implemented via FreeSurfer, version 5.3.0. The current study used post-processed structural data of cortical thickness with the Desikan atlas-based classification (n=68) (Desikan et al 2006). Participants who did not pass visual inspection of T1 images and FreeSurfer quality control (imgincl_t1w_include=1) were excluded from the neuroimaging analysis.

### Statistical analyses

#### Clustering analysis

We capitalized on data-driven clustering techniques to determine subtypes of patients with internalizing disorders based on the five impulsivity dimensions of the UPPS-P model. In line with established guidelines for cluster analyses (Hair 2009) a combination of hierarchical and non-hierarchical procedures was employed. First, we used average silhouette width to determine the optimal number of clusters (Rousseeuw 1987). Second, a hierarchical clustering analysis was conducted to initialize the subsequent K-means clustering. All variables of UPPS-P were scaled, so that each variable contributes equally to the cluster formation. The hierarchical cluster analysis employed Ward’s method and the squared Euclidian distance measure. Third, means of every variable assigned to each cluster were calculated to initialize K-means clustering and cluster assignment was finally fine-tuned by a non-hierarchical K-means cluster analysis.

#### Behavioral and neurobiological (morphological development) characterization

We initially determined phenotypical differences in the domains of anxiety, impulsivity and motivational systems between the identified groups while controlling for age, sex and ethnicity. To characterize neurobiological basis of the phenotype differences linear mixed-effects models (LMMs) were employed to determine differences in cortical thickness between groups. LMMs were employed using *lme4* package (https://cran.r-project.org/web/packages/lme4/index.html). LMMs included random-effects for family ID and acquisition site ID and fixed-effects for dichotomous variable of subject groups (subtype 1, subtype 2 or HC), age, sex, ethnicity, family income, parental years of education, puberty score, body mass index and intracranial volume. False Discovery Rate (FDR, q=0.05) was used for multiple comparisons correction.

#### Neurodevelopmental variations related with anxiety and impulsivity

In addition to the categorical approach, we employed a dimensional approach examining associations between anxiety, impulsivity and cortical thickness in regions exhibiting significant between group differences in the entire sample (n=11878) (similar approach see Xu et al 2020; Xu et al 2021). LMMs were used to regress out the following covariates for neuroimaging variables: family ID and acquisition site ID for random-effects and age, sex, ethnicity, family income, parental years of education, puberty score, body mass index and intracranial volume for fixed-effects. False Discovery Rate (FDR, q=0.05) was used for multiple comparisons correction.

#### Subtype-specific developmental interactions between anxiety and impulsivity

We first used the Pearson correlation coefficient to examine associations between anxiety and impulsivity at baseline. Next, we capitalized on the longitudinal assessments of anxiety and impulsivity to determine whether levels of anxiety at baseline associate with subtype-specific trajectories of impulsivity over the following two years. To this end longitudinal associations between anxiety and impulsivity of the two subtypes were examined using cross-lagged panel models (CLPM) implemented by the *lavaan* package in R (https://cran.r-project.org/web/packages/lavaan/index.html). Anxiety and impulsivity scores were assessed at baseline and 2-year follow-up. The model was estimated by using maximum likelihood estimation. Standardized regression coefficients and p-values are reported. Age, sex, and ethnicity were regressed out as nuisance covariates.

#### Developmental trajectories of psychopathology, suicidality and transition rate to externalizing disorders between internalizing disorder subtypes

Direct comparisons were implemented to compare depressive problems and externalizing psychopathology (measured by the symptom scores on the CBCL) between subtype 1 and 2 at baseline and follow-up. Age, sex, and ethnicity were regressed out as nuisance covariates. False Discovery Rate (FDR, q=0.05) was used for multiple comparisons correction. We also compared the transition rate to externalizing disorders and the prevalence of suicidality (including suicidal ideation, suicide attempt and non-suicidal self-injury) at follow-up of the two subtypes using chi-square test.

#### Subtype-specific academic performance and cognition

As an index of cognitive performance and functioning in daily life we examined differences of academic performance (grades) and cognition between the identified groups at baseline.

## Results

### Sample characteristics

A total n=4066 healthy controls and n=2437 pure internalizing patients at baseline were included in all primary analyses. Demographic information is presented in Table 1.

**Table 1.**
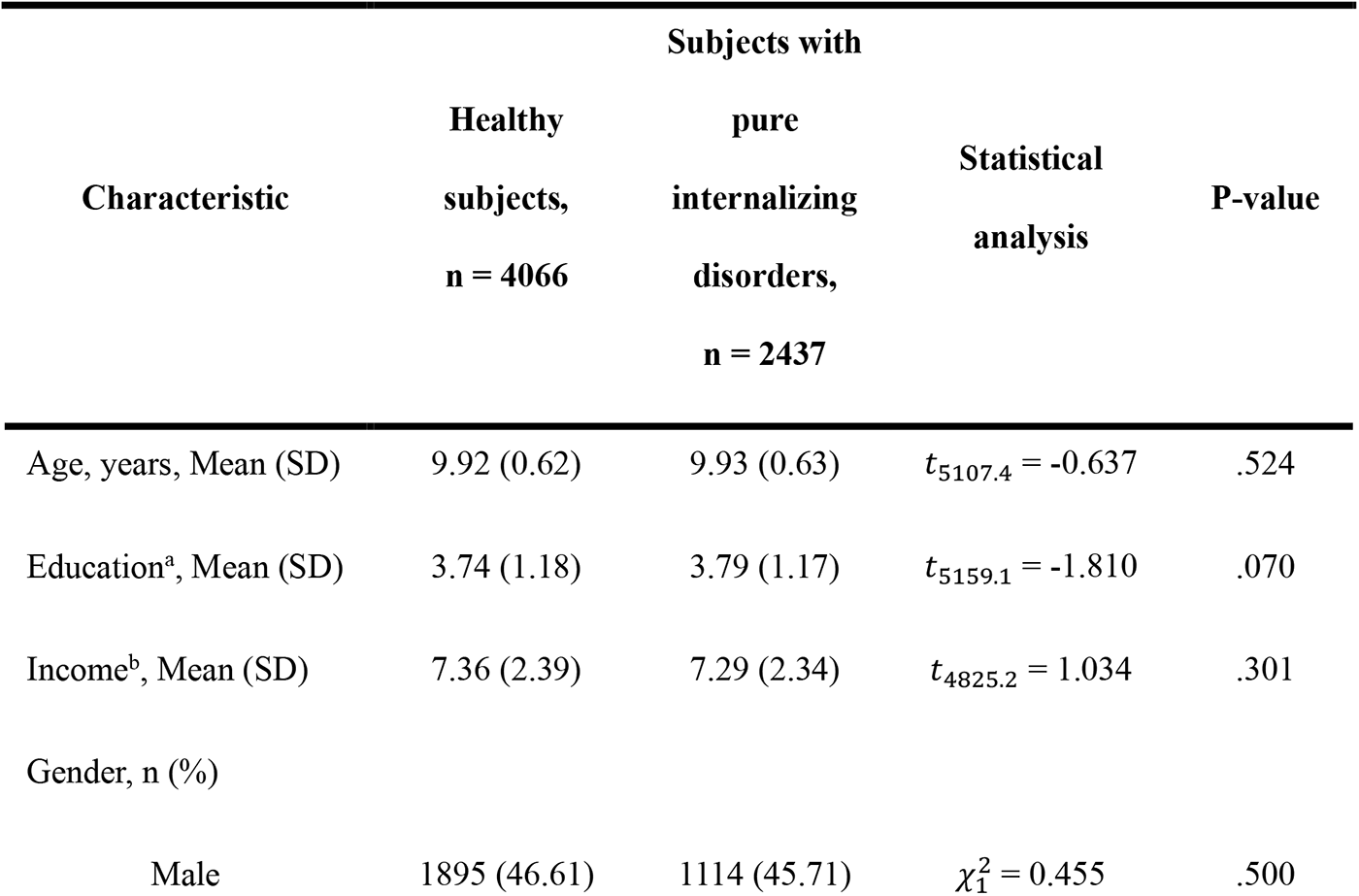
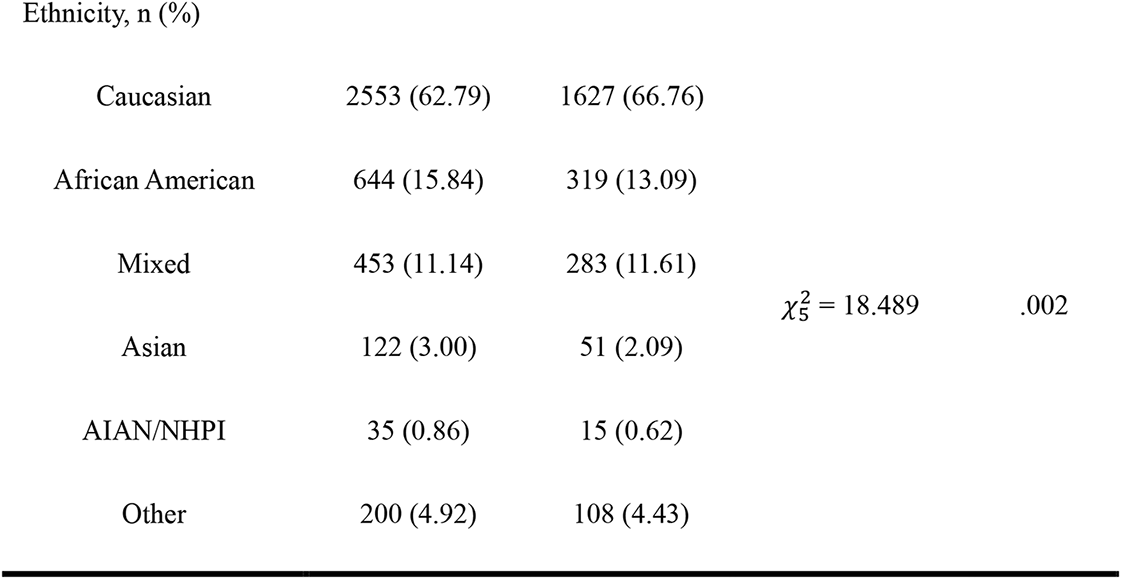
Sample characteristics at baseline. AIAN, American Indian/Alaska Native; NHPI = Native Hawaiian and other Pacific Islander. a: Education of parents was measured by the years of education of the parent with the highest education, categorized as an ordinal variable across five bins (1: < HS Diploma; 2: HS Diploma/GED; 3: Some College; 4: Bachelor; 5: Post Graduate Degree). b: Income was the sum of the annual incomes of both parents, categorized as an ordinal variable across ten bins (1: <$5,000; 2: $5,000-11,999; 3: $12,000-15,999; 4: $16,000-24,999; 5: $25,000-34,999; 6: $35,000-49,999; 7: $50,000-74,999; 8: $75,000-99,999; 9: $100,000-199,999; 10: >$200,000).

### Impulsivity determines distinct internalizing disorder subtypes

Curve of average silhouette width for the clustering indicated a two-cluster solution (Figure S1). Data-driven clustering of the internalizing patients based on the five UPPS-P dimensions revealed two subtypes of pure internalizing patients with high and low impulsivity, respectively (Figure 2A). The subtypes did not differ in terms of the ethnic distribution. However, subtype 2 included slightly more females and they were lower in social economic status. Demographic information and diagnosis of two subtypes are presented in Table S1 and S2, respectively.

**Figure 2.**
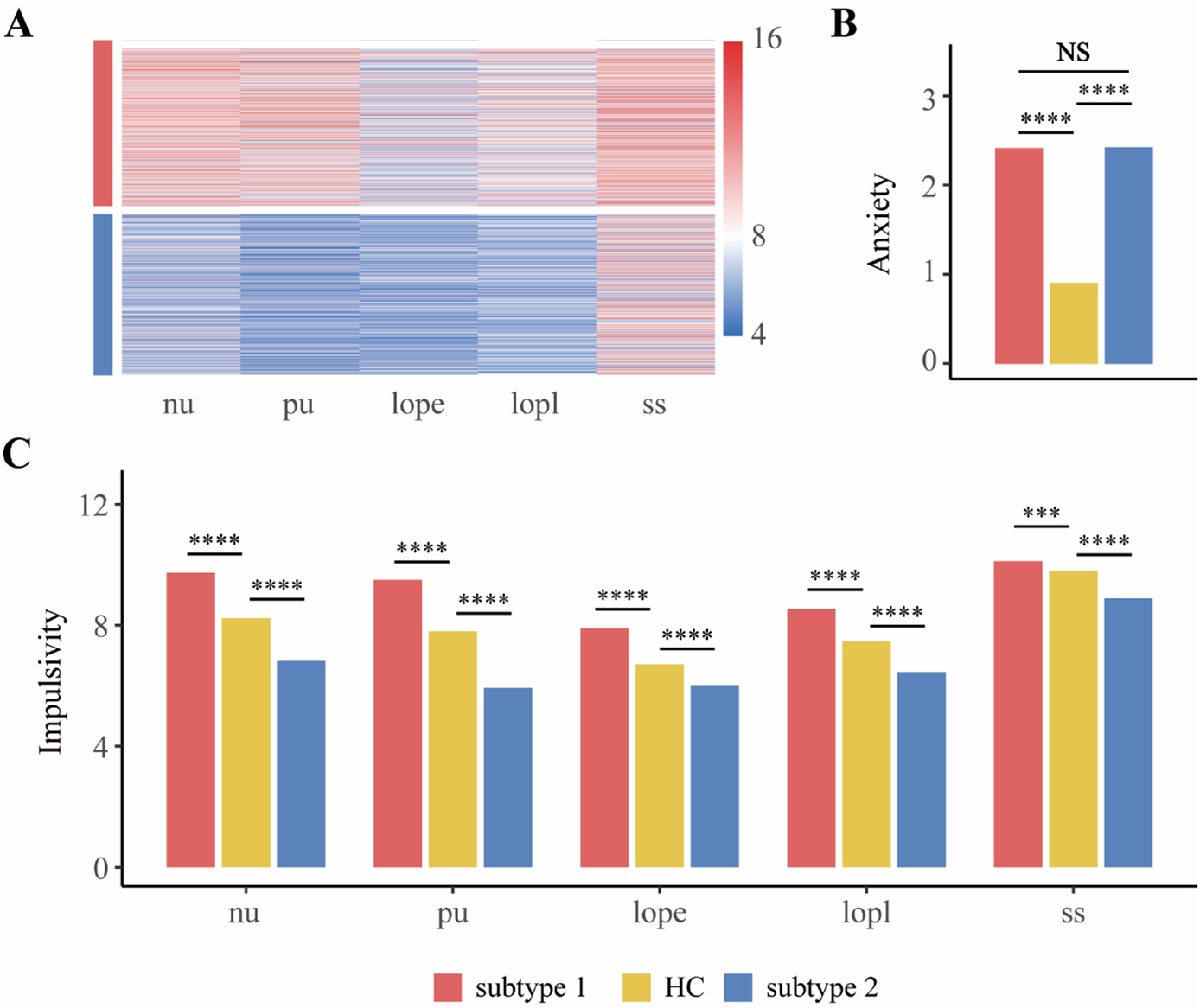
Behavioral differences between subtype 1, subtype 2 in pure internalizing patients and healthy control. (A) Five dimensions of impulsivity (UPPS-P) in two subtypes of internalizing patients (subtype 1 and subtype 2) determined by clustering analysis. (B) Comparisons of anxiety (CBCL-Anxiety Problems). (C) Comparisons of UPPS-P among groups. nu, negative urgency; pu, positive urgency; lope, lack of perseverance; lopl, lack of planning; ss, sensation seeking. HC, healthy control. * p<0.5; ** p<0.01; *** p<0.001; **** p<0.0001; NS, not significant.

We next compared levels of anxiety and impulsivity between subtype 1, subtype 2 and healthy controls (HC) at baseline (Figure 2B, C). Whereas both internalizing groups exhibited comparably exaggerated levels of anxiety relative to controls, subtype 1 exhibited increased while subtype 2 exhibited decreased impulsivity relative to the healthy reference group, respectively. With respect to the motivational systems subtype 1 exhibited increased while subtype 2 exhibited decreased levels of BAS compared to HC (Figure S2). A further longitudinal analysis capitalizing on the 2-year follow up data (Figure S3) revealed that differences in the anxiety, impulsivity and motivational systems remained stable over the follow-up period.

### Internalizing patients with high impulsivity exhibit increased cortical thickness

To determine the neurodevelopmental basis of the phenotypic differences we compared baseline cortical thickness between subtype 1, subtype 2 and HC, respectively. Subtype 1 had significantly thicker cortices than HC in left pars opercularis (t=4.17, p=3.2×10^−5^), pars triangularis (t=2.77, p=5.7 × 10^−3^), caudal middle frontal gyrus (t=3.61, p=3.0×10^−4^), superior frontal gyrus (t=3.23, p=1.2×10^−3^), precentral gyrus (t=3.83, p=1.31×10^−4^), paracentral lobule (t=2.99, p=2.8×10^−3^), inferior temporal gyrus (t=2.85, p=4.3×10^−3^), fusiform gyrus (t=3.35, p=8.1×10^−4^), lingual gyrus (t=2.99, p=2.7×10^−3^) and right inferior temporal gyrus (t=3.31, p=9.6 × 10^−4^) (all significant after FDR correction, Figure 3A), whereas subtype 2 did not display significant differences from HC or subtype 1 (Figure 3B, C).

**Figure 3.**
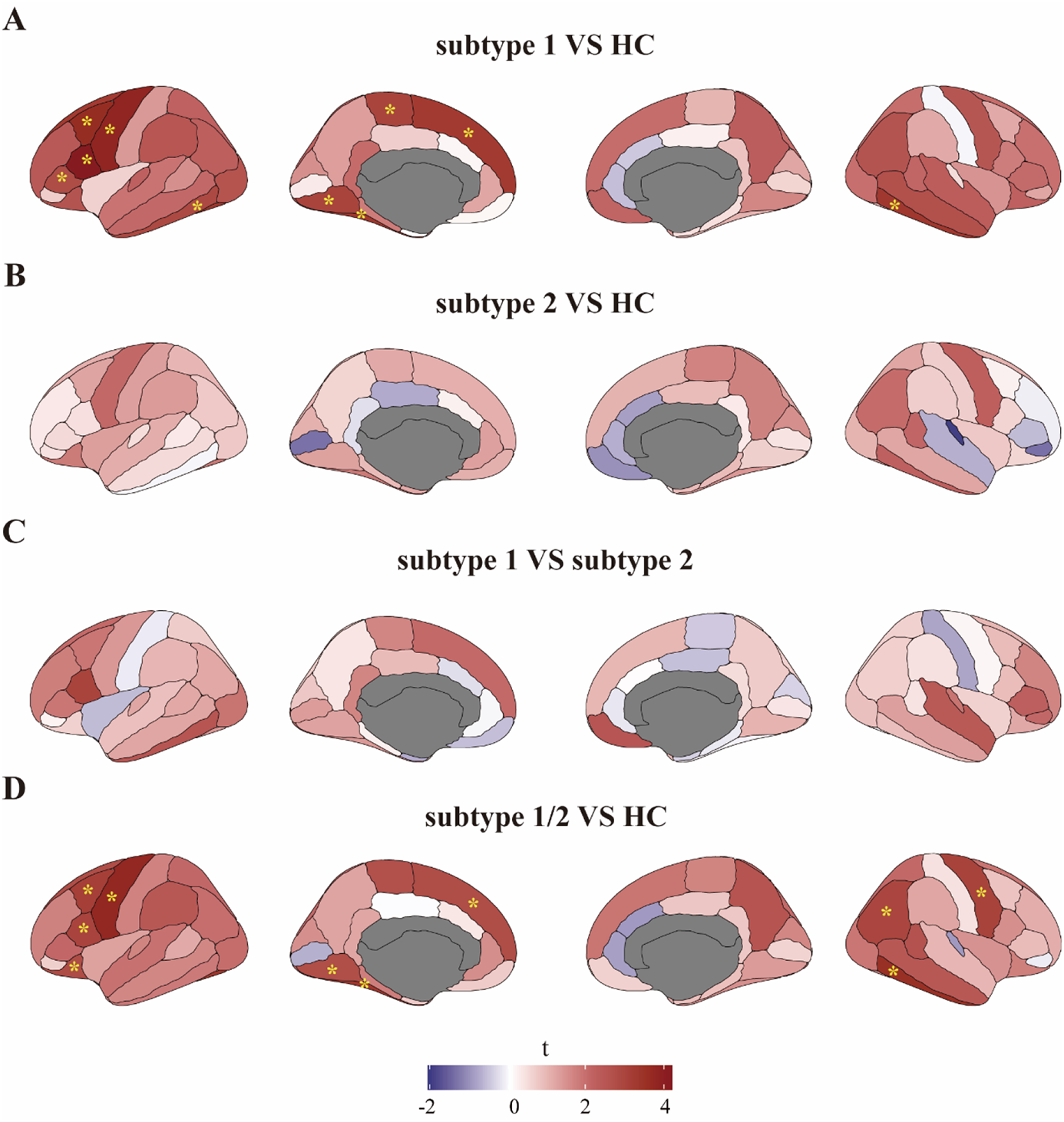
Neurobiological characterization of the subtypes of pure internalizing patients at baseline. (A) Thickness alterations in subtype 1 compared to HC (B) Thickness alterations in subtype 2 compared to HC (C) Thickness alterations in subtype 1 compared to subtype 2. (D) Thickness alterations in all pure internalizing patients (subtype 1 and subtype 2) compared to HC. HC, healthy control. * q<0.05, FDR corrected.

We additionally employed the conventional approach of pooling internalizing patients (subtype 1 and subtype 2) and compared cortical thicknesses between all pure internalizing patients and HC, which revealed similar significant differences compared to the differences between subtype 1 and HC (Figure 3D). Together this suggests that the conventional approach would have revealed cortical thickness alterations in internalizing patients, while the subtype analysis indicated that the differences were clearly and specifically driven by subtype 1, suggesting a neurodevelopmental subtype differentiation on adolescent internalizing disorders.

To investigate the stability of neurodevelopmental alterations between groups, we further compared 2-year follow-up cortical thickness between the subtypes and HC. We found similar differences as those at baseline such that the left pars opercularis (t=3.61, p=3.5 × 10^−4^, FDR corrected), paracentral lobule (t=2.26, p=0.024), fusiform gyrus (t=2.14, p=0.032), precentral gyrus (t=2.10, p=0.036), pars triangularis (t=2.01, p=0.045) and right inferior temporal gyrus (t=2.91, p=0.004) were still thicker in subtype 1 compared to HC (Figure S4A) and subtype 2 did not exhibit any significant differences from HC (Figure S4B). In contrast to the baseline data, the two subtypes differed at follow-up, specifically subtype 1 exhibited thicker left pars opercularis (t=3.78, p=1.6×10^−4)^, right superior temporal gyrus (t=3.58, p=3.6×10^−4^) and middle temporal gyrus (t=3.51, p=4.6×10^−4^) compared to subtype 2 (all significant after FDR correction, see Figure S4C). Examination of the spatial similarity of the thickness distribution by means of employing Pearson correlation analyses between t-maps representing cortical thickness differences of the two subtypes and HC, revealed that the similarity between the subtypes at baseline (r=0.4, p=7×10^−4^, Figure S5A) was decreased at 2-year follow-up (r=0.14, p=0.24, Figure S5B) suggesting that the two subtypes of internalizing disorders become more dissimilar during development indicating divergent neurodevelopmental trajectories. Statistical t-value and p-value of comparisons of cortical thickness are listed in Table S3, S4.

### Higher impulsivity associates with larger cortical thickness of prefrontal cortex

Employing dimensional analysis in the entire sample focusing on regions with significant differences between subtype 1 and HC at baseline with both anxiety and impulsivity scores revealed that larger thickness of left pars triangularis was associated with increased sensation seeking (t=2.49, p=0.013) and larger thicknesses of left pars opercularis and superior frontal gyrus were associated both with increased sensation seeking (t=2.46, p=0.014 and t=2.49, p=0.013, respectively) and positive urgency (t=2.83, p=0.005 and t=2.97, p=0.003, respectively), which all passed FDR correction (Figure S6). Statistical t-value and p-value of the associations are presented in Table S5. Together this analysis suggests that different sub-facets of impulsivity are associated with morphological alterations in different frontal regions.

### Differential longitudinal associations between anxiety and impulsivity in the internalizing subtypes

We investigated the correlation between anxiety and impulsivity at baseline of the two subtypes separately and found two opposing patterns of correlation between them. Anxiety was related with increased lack of perseverance (r=0.10, p=0.0165) in subtype 1 (Figure 4A) but with decreased lack of planning (r=-0.10, p=0.0102) and sensation seeking (r=-0.13, p=0.0009) in subtype 2 (Figure 4B).

**Figure 4.**
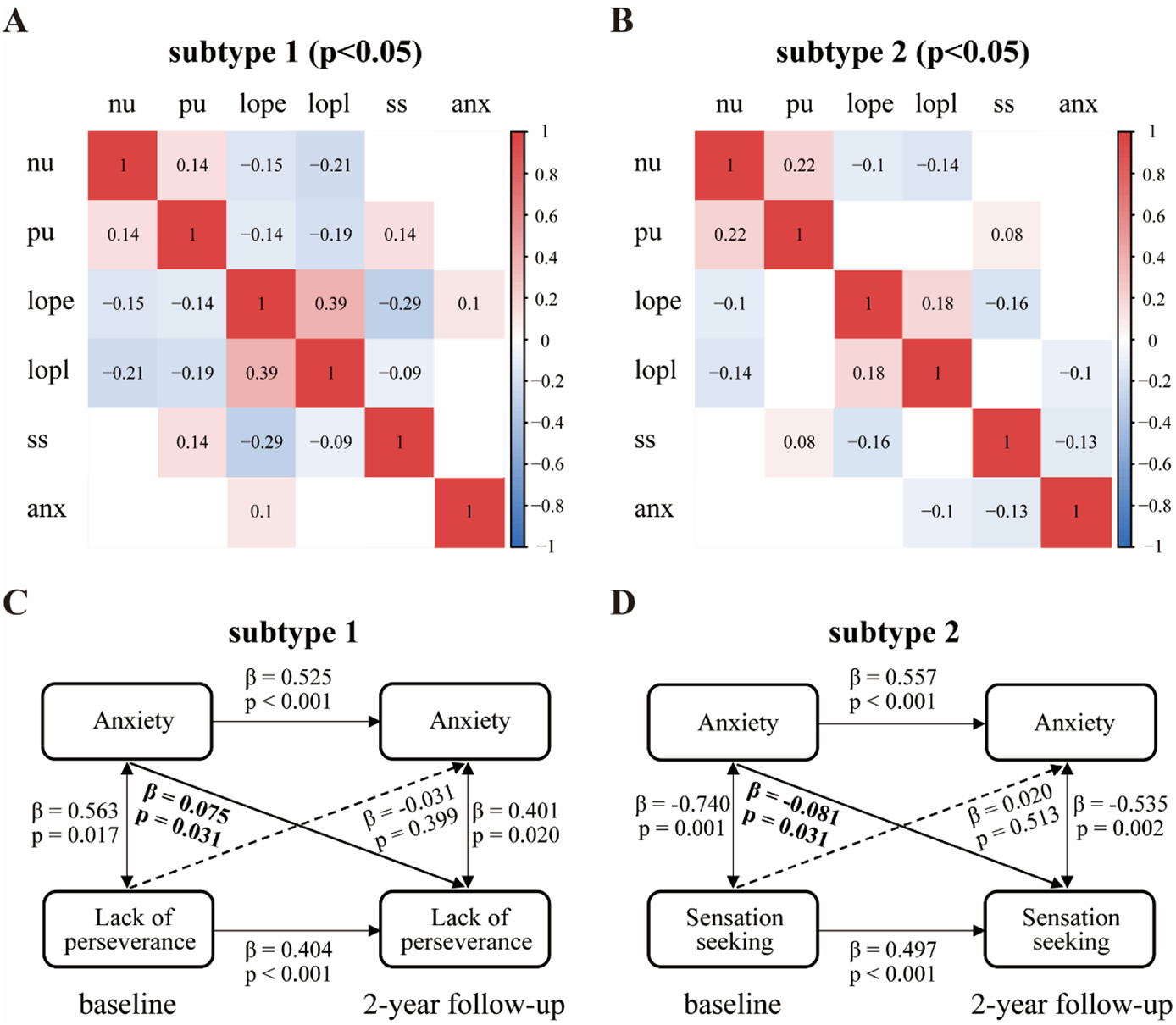
Correlation between anxiety and impulsivity in pure internalizing patients. (A) Baseline anxiety-impulsivity relationship in subtype 1. (B) Baseline anxiety-impulsivity relationship in subtype 2. (C) Longitudinal associations between anxiety and lack of perseverance in subtype 1. (D) Longitudinal associations between anxiety and sensation seeking in subtype 2. Anxiety was measured by CBCL-Anxiety Problem. Impulsivity was measured by sub-facets of UPPS-P. Threshold of significant p-value was 0.05. nu, negative urgency; pu, positive urgency; lope, lack of perseverance; lopl, lack of planning; ss, sensation seeking; anx, anxiety.

We further performed longitudinal association analyses between anxiety and impulsivity over two time points (baseline and 2-year follow-up) of the two subtypes separately by CLPM. In subtype 1 anxiety at baseline was significantly associated with increased lack of perseverance (β = 0.075, p = 0.031) at 2-year follow-up (Figure 4C), while in subtype 2 anxiety at baseline was significantly associated with decreased sensation seeking (β = −0.081, p = 0.031) at 2-year follow-up (Figure 4D), suggesting opposing and impulsivity sub-facet specific associations between baseline anxiety and the trajectories of impulsivity in subtype 1 and subtype 2. Detailed longitudinal associations between anxiety and impulsivity were listed in Table S6.

### Prediction of trajectories: Internalizing patients with high impulsivity exhibit higher psychopathology, higher risk of suicide, higher transition rate to externalizing disorders, poorer cognition and worse academic performance

To further determine distinct psychopathological and neurocognitive profiles between the groups we compared the respective indices between subtype 1 and subtype 2. We found that subtype 1 had higher scores of externalizing problems (e.g., scores of ADHD, ODD and CD in CBCL) compared to subtype 2 at baseline and follow-up, and that subtype 1 had higher depressive symptom load than subtype 2 at 2-year follow-up (Figure 5A). With respect to the clinical predictive utility of the subtypes we found that more patients in subtype 1 developed externalizing disorders (ADHD and ODD) during the 1-year follow-up (Figure 5B) and reported higher prevalence of suicidality, which is mainly driven by suicidal ideation, during the 2-year follow-up compared to subtype 2 (Figure 5C). With respect to functional performance in everyday life we found that subtype 1 had a poorer while subtype 2 had a better academic performance in terms of grades at baseline compared to HC (Figure 5D), and that subtype 2 had a better cognition performance, especially in crystallized intelligence (Figure 5E).

**Figure 5.**
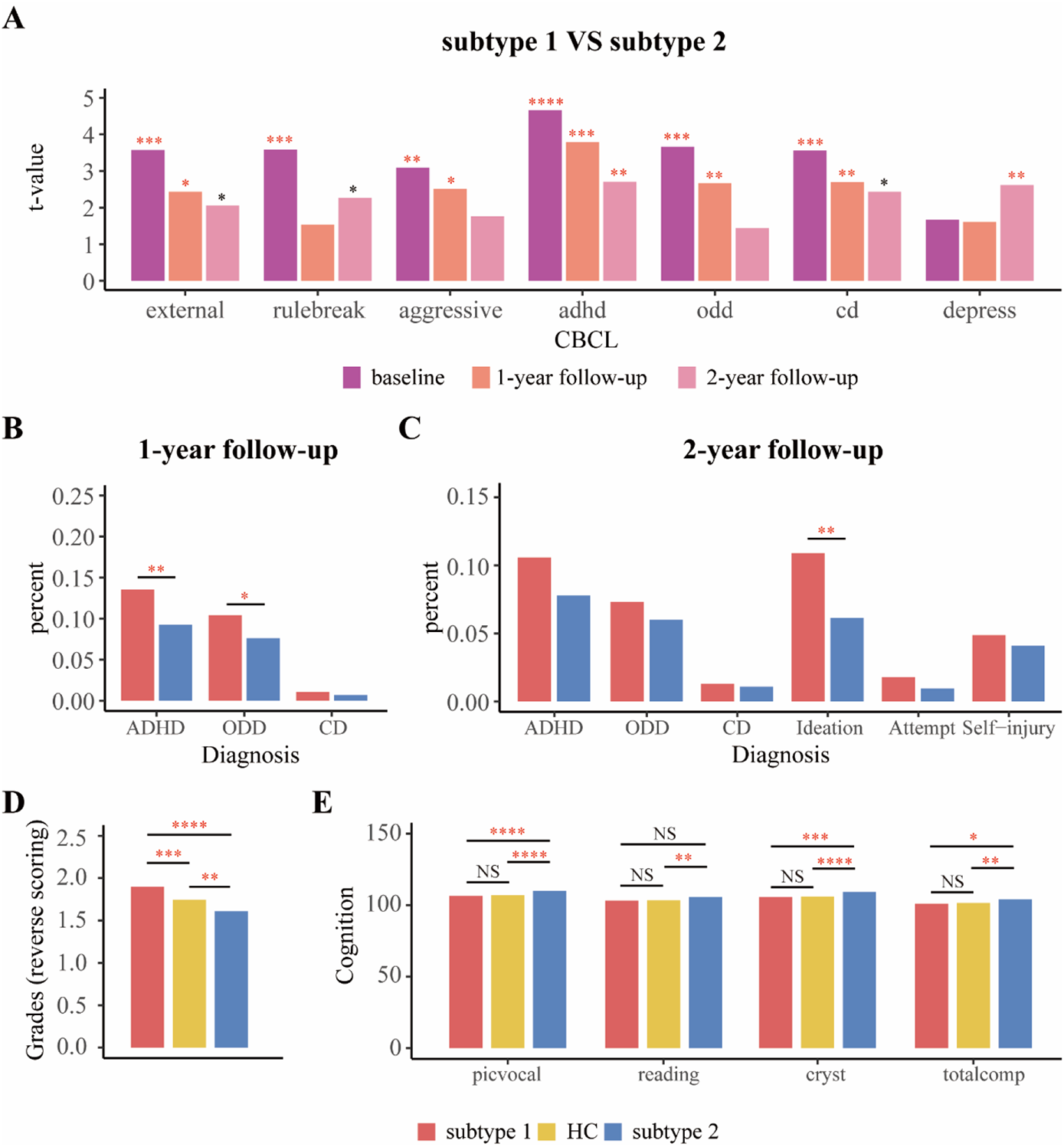
Differences of (A) psychopathology, (B) transition rate to externalizing disorders at 1-year follow-up, (C) transition rate to externalizing disorders and prevalence of suicidality at 2-year follow-up between subgroups in pure internalizing patients, and differences of (D) grades^a^ and (E) cognition at baseline between subgroups in pure internalizing patients and HC. external, Externalizing Problems; rulebreak, Rule-Breaking Behavior; aggressive, Aggressive Behavior; adhd, ADHD Problems; odd, Oppositional Defiant Problems; cd, Conduct Problems; depress, depressive problems; ADHD, Attention-Deficit/Hyperactivity Disorder; ODD, Oppositional Defiant Disorder; CD, Conduct Disorder; Ideation, Suicidal ideation; Attempt, Suicide attempt; Self-injury, Nonsuicidal self-injury; picvoc, picture vocabulary; reading, oral reading recognition; cryst, crystallized intelligence; totalcomp, total intelligence; HC, healthy control. * p<0.05; ** p<0.01; *** p<0.001; **** p<0.0001; NS, not significant. Red asterisks indicate p-values passed FDR correction. a: grades were scored reversely and 1 = excellent, 2= good, 3 = average, 4 = below average, 5 = struggling a lot, and 6 = ungraded.

### Robustness of the clustering results across externalizing disorder comorbidity

Our initial analyses focused on individuals with pure internalizing disorders while excluding subjects with comorbid externalizing disorders to control for a potential influence of comorbid externalizing conditions. To test the robustness of our findings we repeated the analyses including patients with comorbid externalizing conditions. The results of the clustering analysis and between-group differences [including cortical thickness, association between anxiety and impulsivity, level of anxiety and sub-facets of impulsivity, which are shown in Figure S7-9 and Table S7, S8] remained stable, suggesting impulsivity is a transdiagnostic factor, which cuts across all internalizing disorder patients independent of externalizing comorbidity.

## Discussion

Anxiety and impulsivity represent key transdiagnostic dimensions of psychopathology yet their interaction and contribution to emotional disorders in adolescence remains controversial (Jakuszkowiak-Wojten et al 2015) and previous findings of internalizing disorders on the symptomatic and neurobiological level remained inconsistent (Sellnow et al 2020; Kaczkurkin et al 2020; Shackman et al 2013). Combining a large adolescent sample consisting of 2437 internalizing patients aged 9-10 years with a data-driven clustering approach, we identified two distinct subtypes of internalizing patients with comparably elevated levels of anxiety but different levels of impulsivity. Specifically, subtype 1 exhibited elevated, while subtype 2 exhibited lower, levels of impulsivity compared to healthy controls. Our results resonate with recent findings on nonclinical undergraduates or patients with anxiety disorders (Binelli et al 2015; Kashdan and Hofmann 2008; Lipton et al 2016), both characterized by subtypes with similar level of anxiety but different level of impulsivity. The differentiation on the impulsivity dimension was mirrored by different neuroanatomical profiles, opposite relationships between anxiety and impulsivity, different neurocognitive and academic profiles, as well as different clinical trajectories with respect to the conversion to externalizing disorders, prevalence of suicidality, severity of externalizing and depressive problems. Together our results indicate that impulsivity represents a key defining factor for subtypes of young internalizing patients and that the corresponding heterogeneity of internalizing patients may account for the inconsistent findings in the previous literature.

Higher impulsivity in subtype 1 is related to increased thickness in higher order cognitive control networks involving left pars opercularis, pars triangularis and superior frontal gyrus, consistent with previous research (Newman et al 2016; Zhu et al 2020; Whelan et al 2012), and also broadly consistent with a previous study examining impulsivity-brain structural associations in the ABCD study (Owens et al 2020). These brain regions, i.e., ventrolateral and dorsolateral PFC (DLPFC), have been strongly involved in response inhibition (Liddle et al 2001; Zhuang et al 2021). DLPFC has been regarded as a critical region for cognitive control (Chen et al 2018; Miller and Cohen 2001), disruption of which increases choice of immediate rewards (Figner et al 2010). Morphological alterations of PFC in subtype 1 and their lower academic performance in everyday life (Figure 5D) may thus mirror poor cognitive control, maladaptive behaviors under stressful situations and tendency to adopt dysfunctional coping strategies which in the long run may give rise to increased levels of internalizing problems (Lim et al 2021; Reising et al 2018). Furthermore, the intact brain morphology in subtype 2 and the lack of significant correlations between anxiety and cortical thickness in the entire sample suggest that the observed alterations in cortical thickness exhibited subtype 1 are mainly driven by impulsivity rather than anxiety. It is worth noting that impulsivity is not a unitary construct (Strickland and Johnson 2021) and that different dimensions of impulsivity play different roles in disorders on the level of brain structure (Owens et al 2020).

From a neurodevelopmental perspective the observed findings resonate with the dual-system models hypothesizing that the PFC attains functional maturity later than the limbic system and this developmental mismatch results in poor emotion regulation during adolescence (Casey et al 2008; Powers and Casey 2015; Steinberg 2008). The development of thickness of PFC consists of initial childhood increase, following adolescent decrease and adult stabilization (Shaw et al 2008). Thicker PFC in subtype 1 may thus indicate their delayed development, which results in ineffective control over behavioral impulses in response to reward and punishment mediated by the limbic system which in turn may promote engagement in dysfunctional coping strategies under anxiety. Different neurodevelopmental trajectories of the two subtypes are also reflected by the increased thickness differences between them in 2-year follow-up, and by the more dissimilar patterns of cortical thickness alteration compared to HC (t-maps of the two subtypes being non-correlated) at 2-year follow-up compared to baseline.

In addition to alterations in frontal regions, subtype 1 additionally exhibited thicker cortices in occipito- and inferior temporal (fusiform) regions which have been associated with perceptual processes and emotional dysregulation (e.g. Spengler et al 2017; Fusar-Poli et al 2009; Gentili et al 2008) and play a central role in the network related to the pathogenesis of anxiety disorders which also includes the insular cortex, ventromedial prefrontal cortex and amygdala (Xu et al 2021; Strawn et al 2014). In line with the functional characterization, greater thickness of these regions has been previously observed in pediatric generalized anxiety disorder and has been associated with fear learning, fear extinction, and deficits in the regulation of the amygdala (Strawn et al 2014), suggesting that developmental delay within these regions may promote a lack of adaptive responses during stressful and anxiogenic situations.

In subtype 1 higher anxiety at baseline preceded increasing impulsivity (lack of planning) over the subsequent two years, while baseline anxiety in subtype 2 correlated negatively with impulsivity (sensation seeking) at two-year follow-up. These two distinct subtypes may harmonize the previous controversial findings. On the one hand, anxiety might serve a critical defensive function for avoiding potential danger (Lee et al 2006; Taylor et al 2008), which has been considered to be inversely related to impulsivity (Apter et al 1993; Taylor et al 2008). This proposed pattern mirrors the findings in subtype 2. On the other hand, recent studies found that patients with a comorbid anxiety disorder had higher impulsivity than those without an anxiety disorder (Del Carlo et al 2012; Ferreira-Garcia et al 2019; Perugi et al 2011; Summerfeldt et al 2004; Taylor et al 2008), which indicates that anxiety can be positively related with impulsivity (Cosi et al 2011; Favaloroa and Moustafab 2020; Jakuszkowiak-Wojten et al 2015; Moustafa et al 2017; Yu et al 2020). This positive relationship appears to be a consequence of the greater salience of immediacy, overestimation of the value of immediate rewards and greater motivation to respond to immediacy in anxious people (Xia et al 2017). This corresponds to the characteristics that we identified in subtype 1.

The distinct anxiety-impulsivity relationship may suggest different strategies adopted by the two subtypes of internalizing patients to cope with anxiety. Subtype 1 may adopt dysfunctional coping like avoiding behaviors, which likely reduces anxiety temporarily but will lead to a maintenance of anxiety in future similar situations (Thomasson and Psouni 2010). Anxiety is believed to play a role in externalizing disorders characterized by impulsive behavior, particularly internet addiction (Bargeron and Hormes 2017; Peterka-Bonetta et al 2019), gambling (Devos et al 2020), alcohol and substance use (Adams et al 2019; Davis et al 2020; Zhou et al 2019). These impulsive behaviors serve to reduce the underlying emotional tension and provide immediate rewards (Tice et al 2001). Moreover, we also found that subtype 1 exhibits significantly higher BAS than controls while subtype 2 has significantly lower BAS than controls. The BAS refers to biologically based behavioral tendencies in response to rewarding stimuli and has been linked with impulsivity (Vollrath and Torgersen 2000). The high BAS scores of subtype 1 may thus reflect sensitivity to rewards, which may impede coping with their anxiety, while the lower impulsivity and BAS in subtype 2 suggest that they are less sensitive to distractions by rewards and thus more likely to resolve problems that caused their anxiety.

Psychopathological markers like suicidal ideation, depressive and externalizing symptoms at baseline and follow-up were all increased in subtype 1, with suicidal ideation and depression being important predictors of suicide in adolescents (Hubers et al 2018; Hawton et al 2013). Previous studies (Lipton et al 2016; Askenazy et al 2003) also reported an association between increased impulsivity and suicidality in internalizing patients. Suicidality encompasses suicidal ideation (Tsypes et al 2019), suicide attempt (Clark et al 2011; Dombrovski et al 2010) and nonsuicidal self-injury (Lutz et al 2021; Wilkinson et al 2018), and in the present study particularly suicidal ideation was found to contribute to the differences between subtypes which may reflect a high clinical relevance given that suicidal ideation has been associated with actual suicide attempts (Chapman et al 2015; Hubers et al 2018). Impulsivity, anxiety and depression are often immediate suicide risk factors that are potentially modifiable if recognized and treated urgently with effective medications and watchful support (Fawcett 2001). In addition, subtype 1 exhibited further cognitive problems, possible reflecting deficient or retarded neurocognitive and functional development, including lower academic grades in daily life. Notably subtype 2 exhibited higher crystallized intelligence and academic performance as compared to both other groups, potentially reflecting a neurocognitive compensation allowing adaptation despite elevated levels of anxiety. Together, subtype 1 may represent a group with a strongly increased psychopathological risk and a risk group for future behavioral and neurodevelopmental dysregulations. Therefore, early interventions for this subgroup, such as Cognitive Behavioral Therapy (CBT), which has been shown to alleviate anxiety in internalizing patients with high impulsivity (Subotic-Kerry et al 2016), are urgently needed.

### Strengths and Limitations

One strength of the present study is the fully data-driven determination of different subtypes in a large longitudinal cohort, which revealed subgroups of internalizing patients with distinct neuroanatomical, neurocognitive and psychopathological profiles at baseline and further diverging trajectories over the follow-up period. Moreover, the data-driven clustering results, neurodevelopmental profiles and distinct anxiety-impulsivity relationships in subtype 1 and subtype 2 remained stable using both pure internalizing patients and internalizing patients comorbid with externalizing disorders, indicating that anxiety-impulsivity relationships represent a transdiagnostic factor that cuts across diagnostic categories and comorbidity externalizing conditions in internalizing disorders. Although there were no differences in the gender distribution of internalizing patients and healthy controls in general, subtype 2 had a higher ratio females, suggesting the potential contribution of sex-differences which may be further explored in future studies. The findings have to be considered in the context of limitations, including the focus on brain morphological data while future studies should examine alterations on the level of functional activity and networks which may allow to characterized brain functional alterations underlying enhanced anxiety in subtype 2.

## Data Availability

All data produced are available online at https://nda.nih.gov/study.html?id=901

## Acknowledgments and Disclosures

JZ was supported by Shanghai Municipal Science and Technology Major Project (No.2018SHZDZX01) and ZJLab and NSFC 61973086. JF was supported by the 111 Project (No. B18015), the key project of Shanghai Science and Technology (No. 16JC1420402), National Key R&D Program of China (No. 2018YFC1312900), National Natural Science Foundation of China (NSFC 91630314). BB is supported by the National Key Research and Development Program of China (2018YFA0701400).

Data used in the preparation of this article were obtained from the Adolescent Brain Cognitive Development (ABCD) study (https://abcdstudy.org), held in the National Institute of Mental Health Data Archive. This is a multisite, longitudinal study designed to recruit more than 11,000 children aged 9 to 10 and follow them over 10 years into early adulthood. The ABCD study is supported by National Institutes of Health and additional federal partners under Grant U01DA041089, U24DA041123, U01DA041117, U01DA041022, U01DA041148, U01DA041106, U01DA041028, U01DA041048, U24DA041147, U01DA041156, U01DA041134, U01DA041174, U01DA041120, U01DA041093, U01DA041025, U01DA050989, U01DA051039, U01DA051016, U01DA051037, U01DA050987, U01DA051018, U01DA051038 and U01DA050988. A full list of supporters is available at https://abcdstudy.org/federal-partners/. A listing of participating sites and a complete listing of the study investigators can be found at https://abcdstudy.org/scientists/workgroups/. The ABCD Research Consortium investigators designed and implemented the study and/or provided data but did not necessarily participate in analysis or writing of this report. This manuscript reflects the views of the authors and may not reflect the opinions or views of the National Institutes of Health or ABCD Research Consortium investigators. The ABCD study data repository grows and changes over time. The ABCD study data used in this report came from Data Release 3.0 (https://nda.nih.gov/study.html?id=901).

BJS consults for Cambridge Cognition, Greenfield BioVentures, and Cassava Sciences. TWR consults for Cambridge Cognition, Shionogi, Heptares, Takeda, Arcadia, and Greenfield Bioventures; and receives royalties from Cambridge Cognition and research grants from Shionogi and GlaxoSmithKline. All other authors report no biomedical financial interests or potential conflicts of interest.

