## Supplemental Table S1-S2, Supplemental Figure S1-S9 for "Anxiety-impulsivity subtypes in adolescent internalizing disorder are characterized by distinguishable neurodevelopmental, neurocognitive and clinical trajectory signatures"

### Supplementary information

| Characteristic | Subtype 1, n = 1203 | Subtype 2, n = 1227 | Statistical analysis | P-value |
|---|---|---|---|---|
| Age, years, Mean (SD) | 9.89 (0.63) | 9.96 (0.62) | $t_{2425.4}$ = -2.950 | .003 |
| Education[a], Mean (SD) | 3.69 (1.20) | 3.89 (1.13) | $t_{2140.6}$ = -4.253 | < .001 |
| Income[b], Mean (SD) | 7.17 (2.39) | 7.41 (2.27) | $t_{2221.8}$ = -2.431 | .015 |
| Gender, n (%) | | | | |
| Male | 618 (51.37) | 491 (40.02) | $\chi_1^2$ = 31.114 | < .001 |
| Ethnicity, n (%) | | | | |
| Caucasian | 773 (64.26) | 850 (69.27) | | |
| African American | 164 (13.63) | 153 (12.47) | | |
| Mixed | 149 (12.39) | 134 (10.92) | | |
| Asian | 27 (2.24) | 23 (1.87) | $\chi_5^2$ = 10.924 | .053 |
| AIAN/NHPI | 5 (0.42) | 10 (0.81) | | |
| Other | 65 (5.40) | 43 (3.50) | | |

Table S1 Demographic information of two subtypes in pure internalizing patients at baseline. AIAN, American Indian/Alaska Native; NHPI = Native Hawaiian and other Pacific Islander.

a: Education of parents was measured by the years of education of the parent with the highest education, categorized as an ordinal variable across five bins (1: < HS Diploma; 2: HS Diploma/GED; 3: Some College; 4: Bachelor; 5: Post Graduate Degree).

b: Income was the sum of the annual incomes of both parents, categorized as an ordinal variable across ten bins (1: <$5,000; 2: $5,000-11,999; 3: $12,000-15,999; 4: $16,000-24,999; 5: $25,000-34,999; 6: $35,000-49,999; 7: $50,000-74,999; 8: $75,000-99,999; 9: $100,000-199,999; 10: >$200,000).

| Diagnosis, n (%) | Subtype 1, n = 1203 | Subtype 2, n = 1227 | Statistical analysis | P-value |
|---|---|---|---|---|
| Depression | 155 (12.88) | 110 (8.96) | $\chi_1^2$ = 9.205 | .002 |
| Panic disorder | 15 (1.25) | 19 (1.55) | $\chi_1^2$ = 0.212 | .645 |
| Agoraphobia | 10 (0.83) | 11 (0.90) | $\chi_1^2$ < 0.001 | 1 |
| Separation anxiety disorder | 331 (27.51) | 387 (31.54) | $\chi_1^2$ = 4.538 | .033 |
| Social anxiety disorder | 136 (11.31) | 152 (12.39) | $\chi_1^2$ = 0.582 | .445 |

| | | | | |
|---|---|---|---|---|
| Specific phobia | 833 (69.24) | 849 (69.19) | $\chi_1^2 = 0$ | 1 |
| Generalized anxiety disorder | 98 (8.15) | 111 (9.05) | $\chi_1^2 = 0.517$ | .472 |
| Posttraumatic stress disorder | 85 (7.07) | 60 (4.89) | $\chi_1^2 = 4.744$ | .029 |
| Obsessive-compulsive disorder | 136 (11.31) | 138 (11.25) | $\chi_1^2 < 0.001$ | 1 |
| Neurodevelopmental disorder | 305 (25.35) | 306 (24.94) | $\chi_1^2 = 0.036$ | .850 |
| Eating disorder | 136 (11.31) | 122 (9.94) | $\chi_1^2 = 1.048$ | .306 |
| Homicidal problems | 4 (0.33) | 2 (0.16) | $\chi_1^2 = 0.187$ | .665 |
| Sleeping problems | 146 (12.14) | 101 (8.23) | $\chi_1^2 = 9.720$ | .002 |
| Suicidal ideation | 98 (8.15) | 64 (5.22) | $\chi_1^2 = 7.918$ | .005 |
| Suicide attempt | 13 (1.08) | 6 (0.49) | $\chi_1^2 = 2.031$ | .154 |
| Nonsuicidal self-injury | 50 (4.16) | 41 (3.34) | $\chi_1^2 = 0.904$ | .342 |

Table S2 Diagnosis of two subtypes in pure internalizing patients at baseline.

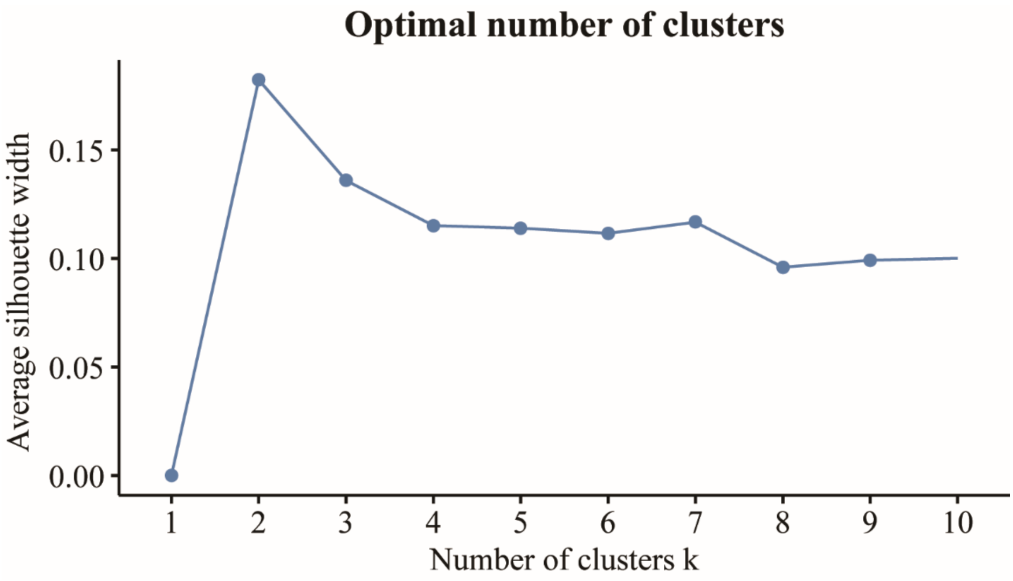


Figure S1 Curve of average silhouette width in the clustering analysis of pure internalizing patients.

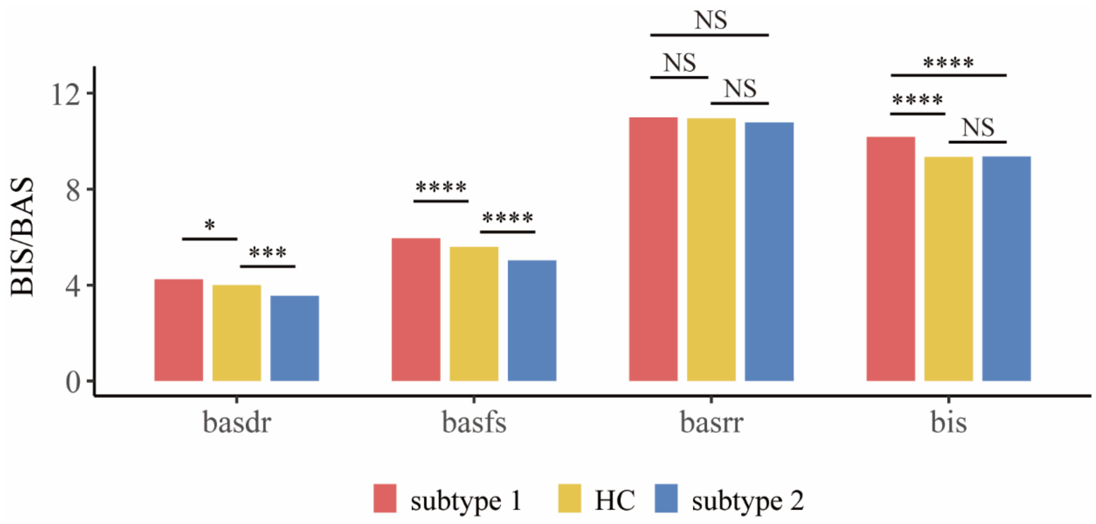


Figure S2 Differences of BIS/BAS between groups in pure internalizing patients at baseline. basdr, behavioral activation: Drive; basfs, behavioral activation: Fun seeking; basrr, behavioral activation: Reward Responsiveness; bis, behavioral inhibition; HC, healthy control. * p<0.5; ** p<0.01; *** p<0.001; **** p<0.0001; NS, not significant.

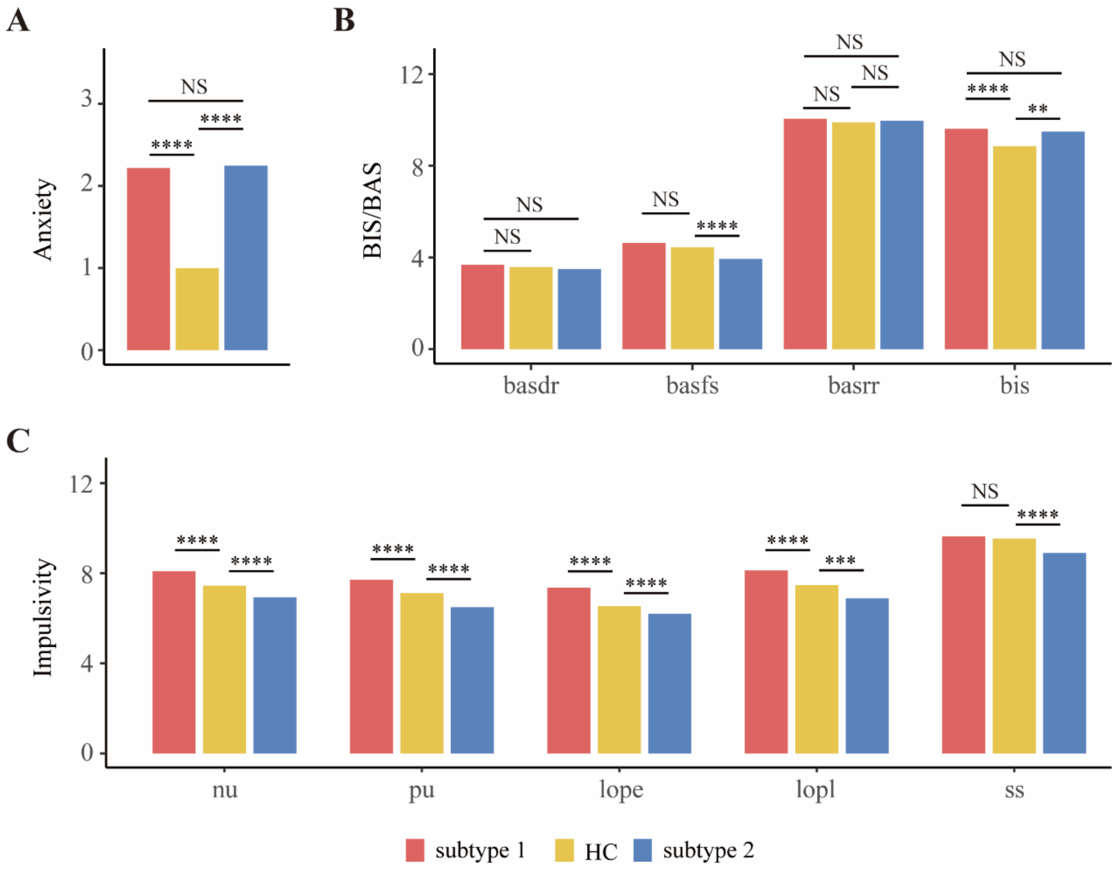


Figure S3 Behavioral differences between groups in pure internalizing patients at 2-year follow-up. (A) Comparisons of anxiety (CBCL-Anxiety Problems) among groups. (B) Comparisons of UPPS-P among groups. (C) Comparisons of BIS/BAS among groups. basdr, behavioral activation: Drive; basfs, behavioral activation: Fun seeking; basrr, behavioral activation: Reward Responsiveness; bis, behavioral inhibition; nu, negative urgency; pu, positive urgency; lope, lack of perseverance; lopl, lack of planning; ss, sensation seeking; HC, healthy control. * p<0.5; ** p<0.01; *** p<0.001; **** p<0.0001; NS, not significant.

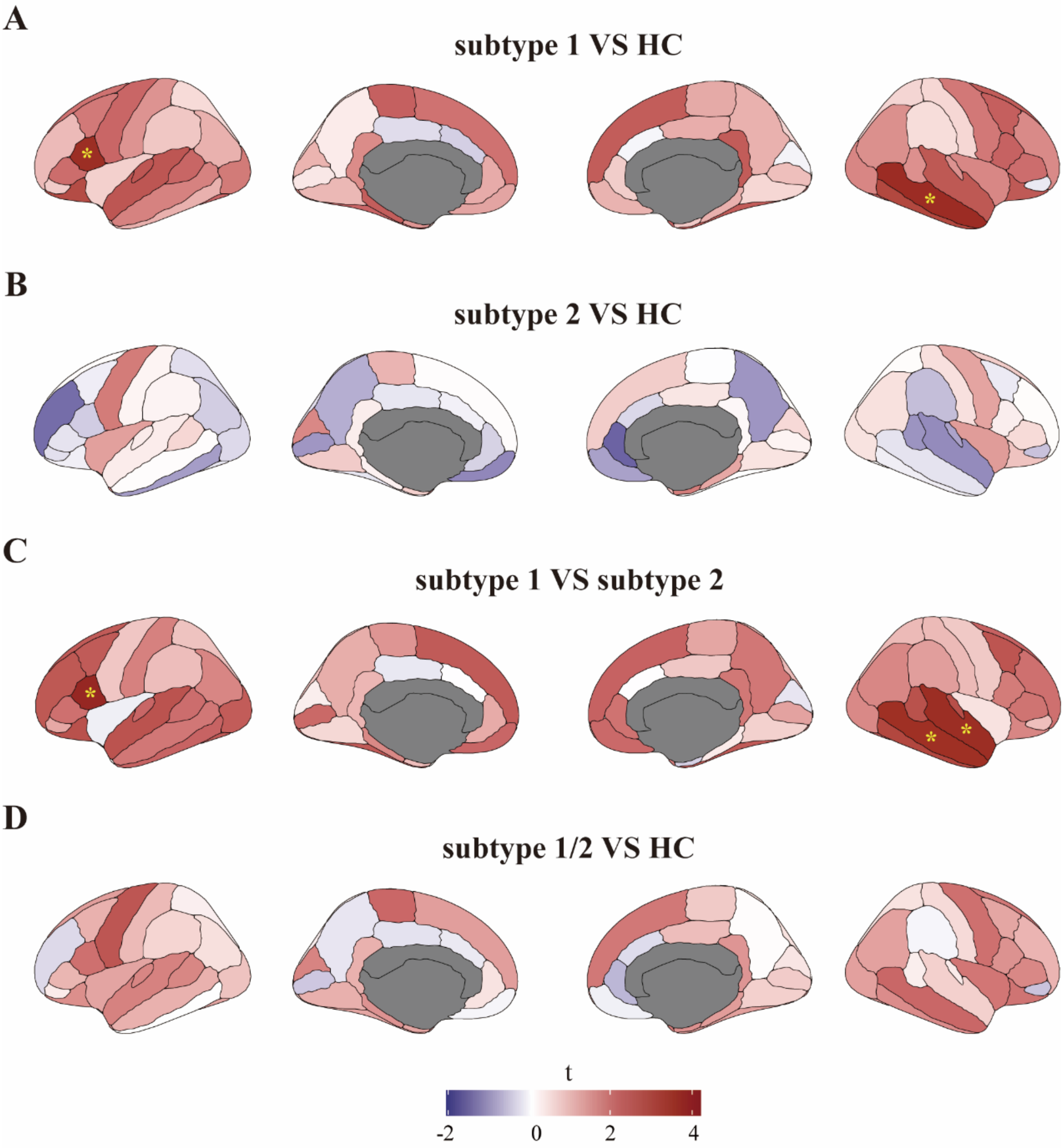


Figure S4 Neurobiological characterization of the subtypes of pure internalizing patients at 2-year follow-up. (A) Thickness alterations in subtype 1 compared to HC. (B) Thickness alterations in subtype 2 compared to HC. (C) Thickness alterations in subtype 1 compared to subtype 2. (D) Thickness alterations in all pure internalizing patients (subtype 1 and subtype 2) compared to HC. HC, healthy control. * q<0.05, FDR corrected.

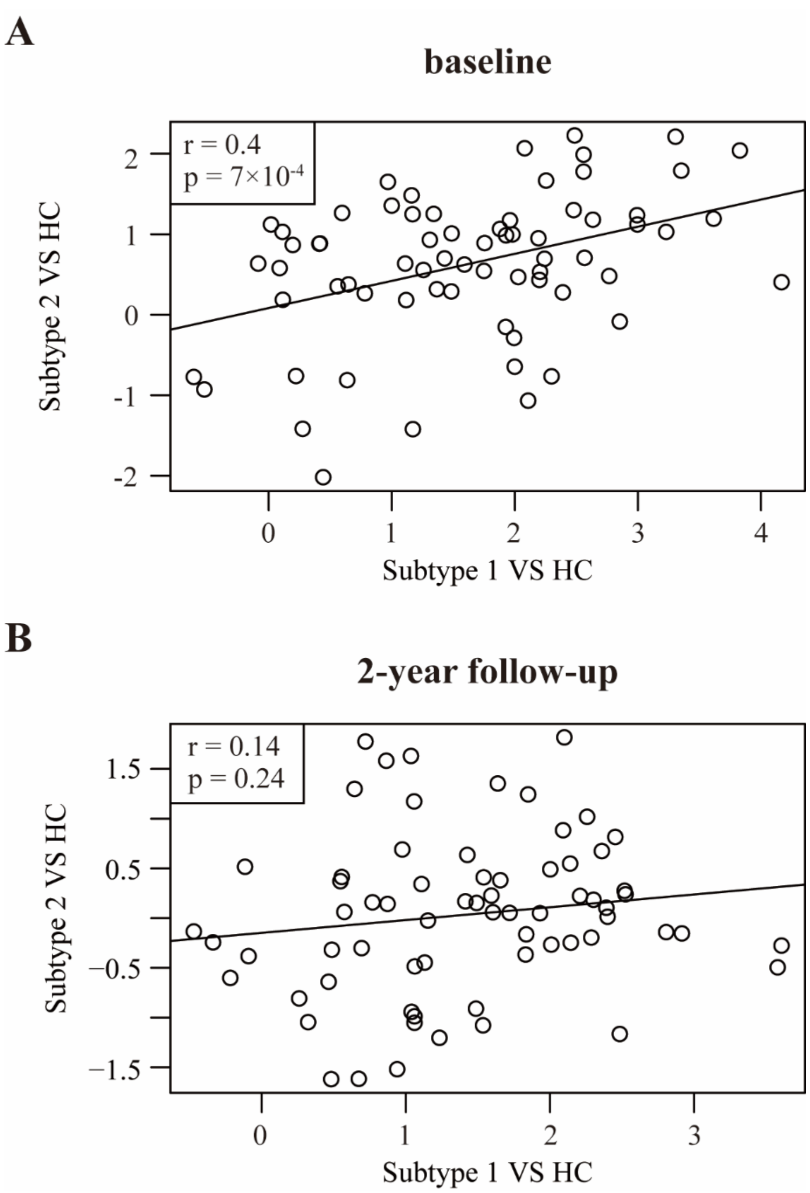


Figure S5 Correlation between t-maps representing cortical thickness differences of the two subtypes and HC at (A) baseline and (B) 2-year follow-up. Each circle represents t-value of cortical thickness difference in a brain region.

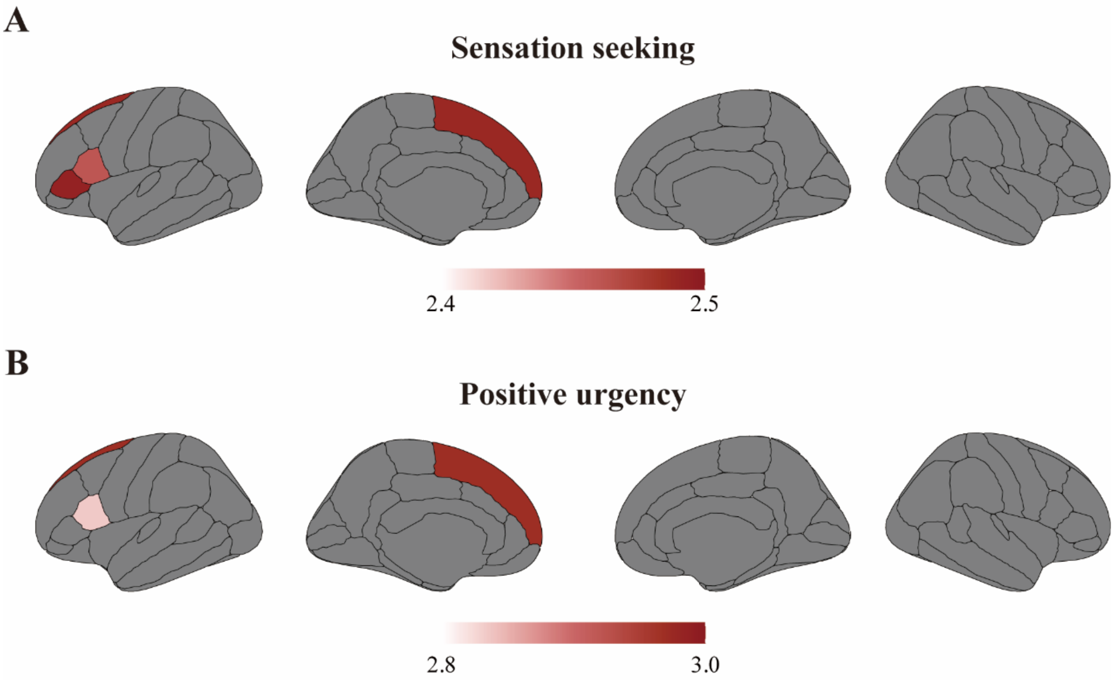


Figure S6 Correlation between impulsivity and cortical thickness in the entire sample. Regions with significant difference between subtype 1 (in pure internalizing patients) and HC at baseline were examined. (A) Correlation between sensation seeking and cortical thickness at baseline. (B) Correlation between positive urgency and cortical thickness at baseline. All passed FDR correction at the threshold of 0.05.

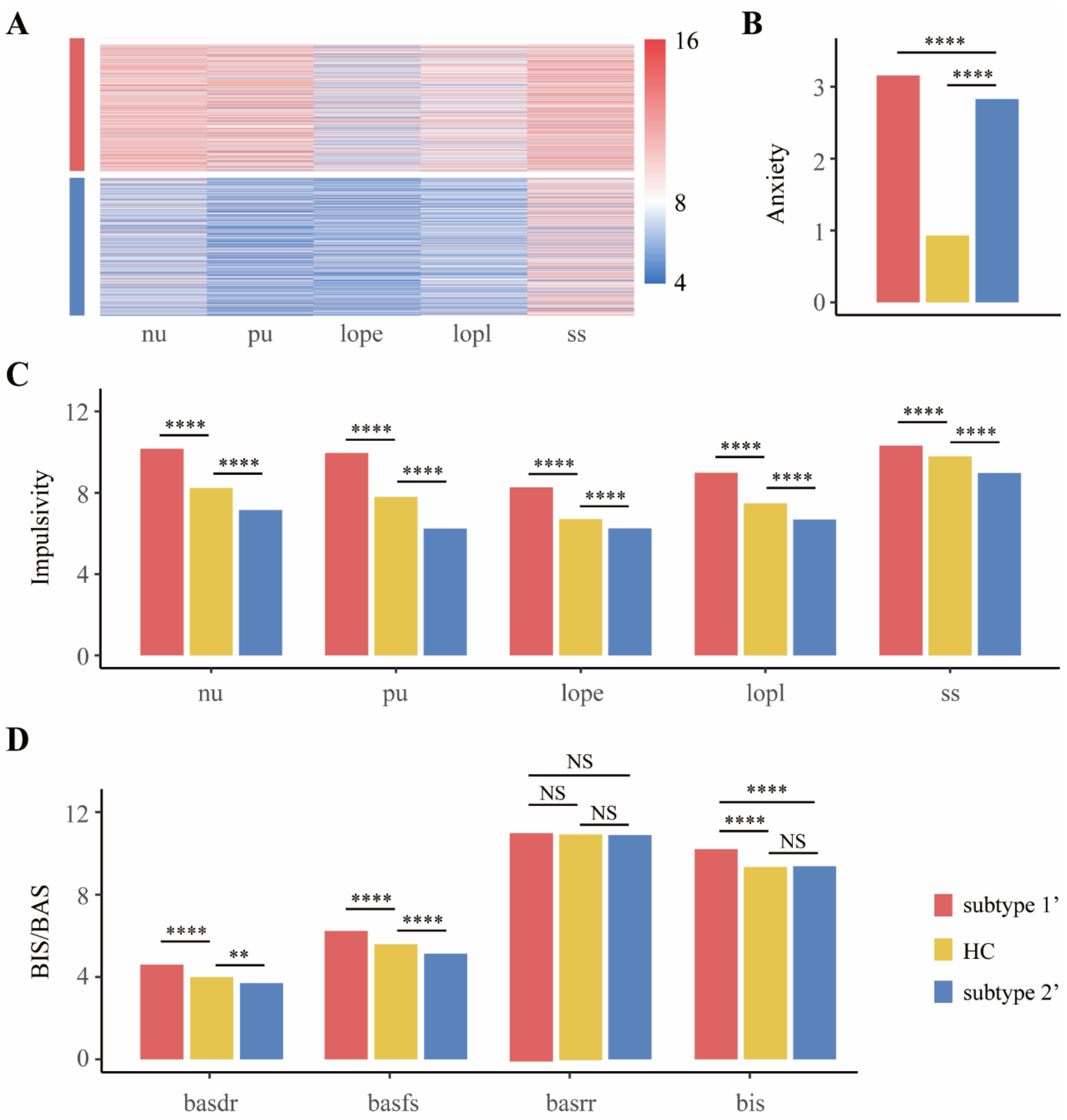


Figure S7 Classification and behavioral differences between groups in all internalizing patients at baseline. (A) Five dimensions of impulsivity (UPPS-P) in two subtypes of internalizing patients (subtype 1’ and subtype 2’) determined by clustering analysis. (B) Comparisons of anxiety (CBCL-Anxiety Problems) among groups. (C) Comparisons of UPPS-P among groups. (D) Comparisons of BIS/BAS among groups. nu, negative urgency; pu, positive urgency; lope, lack of perseverance; lopl, lack of planning; ss, sensation seeking; basdr, behavioral activation: Drive; basfs, behavioral activation: Fun seeking; basrr, behavioral activation: Reward Responsiveness; bis, behavioral inhibition; HC, healthy control. * p<0.5; ** p<0.01; *** p<0.001; **** p<0.0001; NS, not significant.

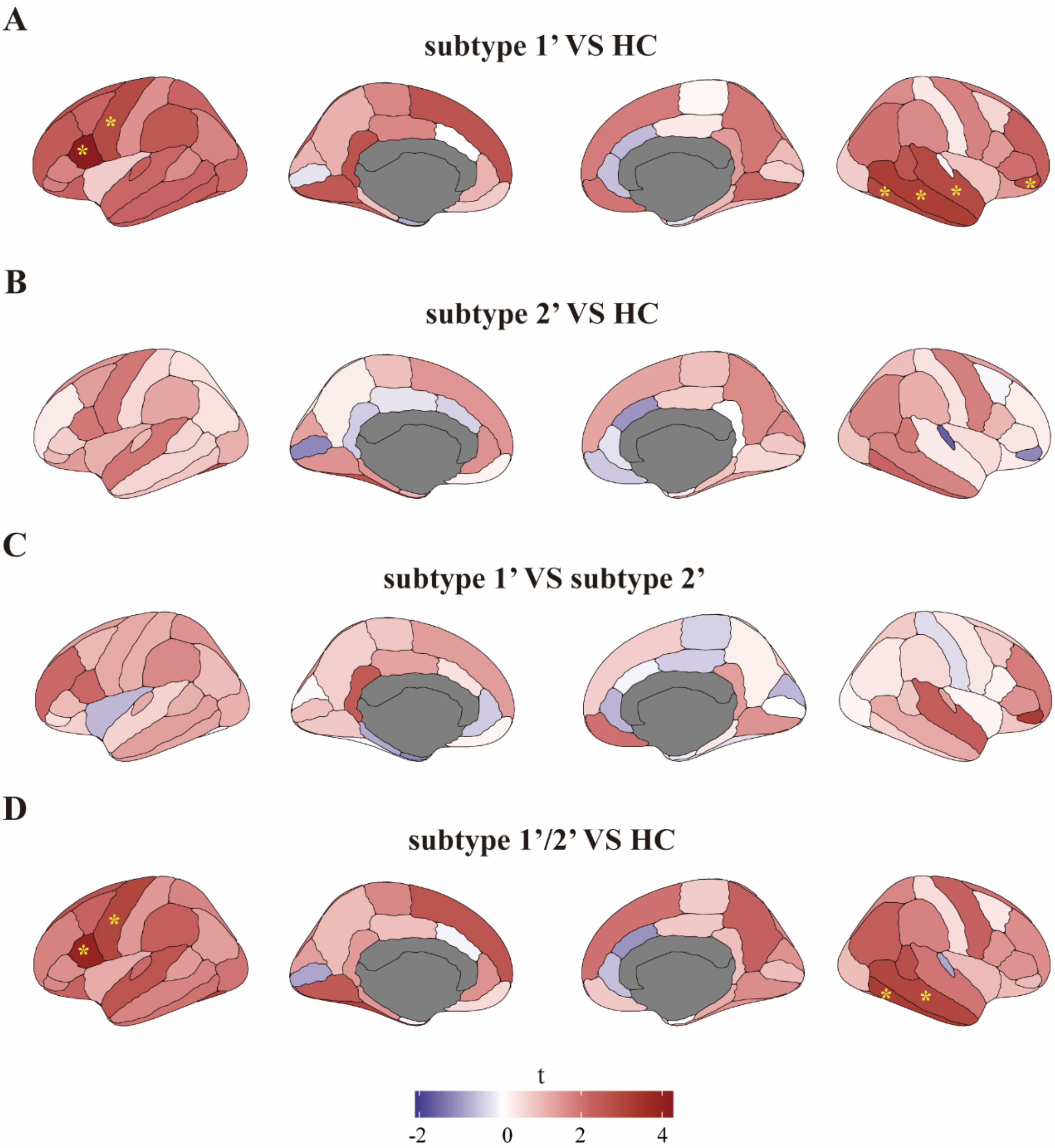


Figure S8 Differences of cortical thickness between groups in all internalizing patients at baseline. (A) Thickness alterations in subtype 1’ compared to HC. (B) Thickness alterations in subtype 2’ compared to HC. (C) Thickness alterations in subtype 1’ compared to subtype 2’. (D) Thickness alterations in all internalizing patients (subtype 1’ and subtype 2’) compared to HC. HC, healthy control. * q<0.05, FDR corrected.

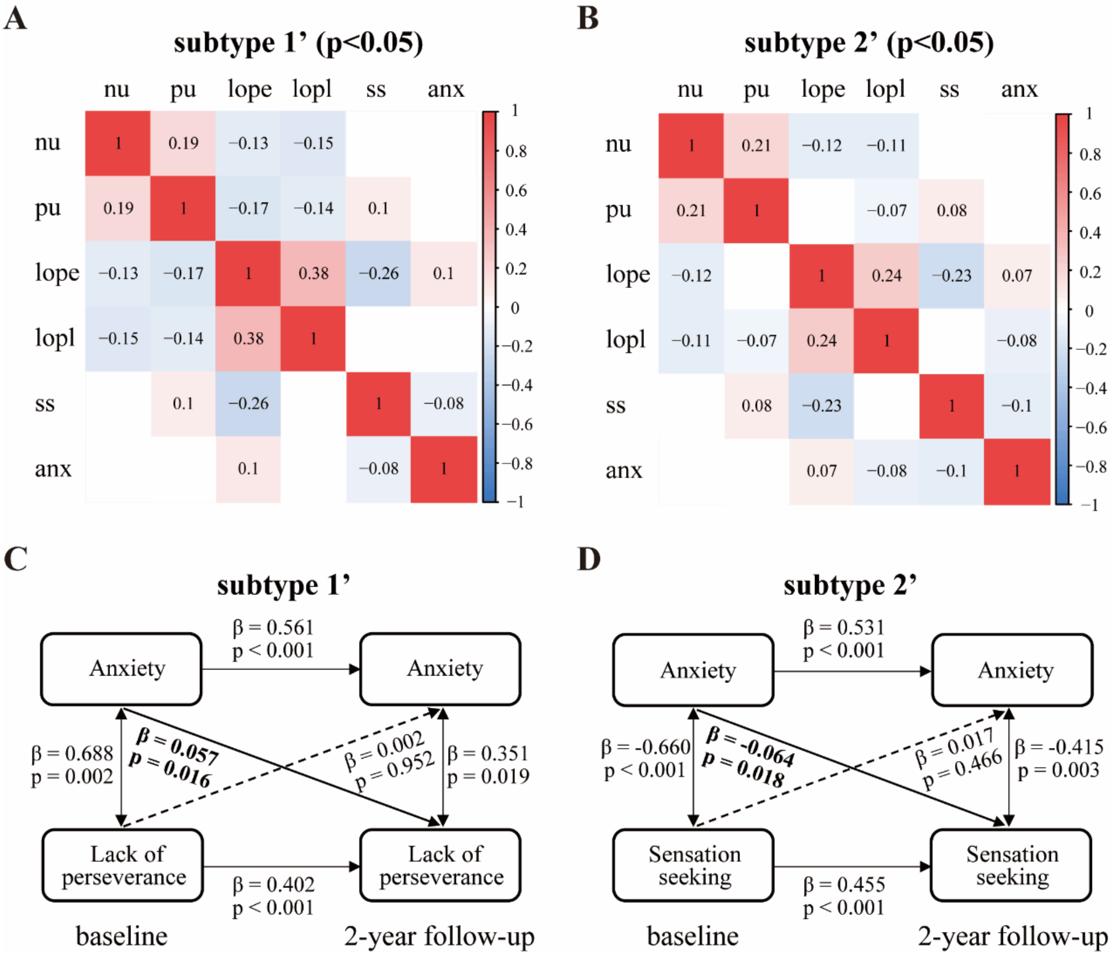


Figure S9 Correlation between anxiety and impulsivity in all internalizing patients. (A) Baseline anxiety-impulsivity relationship in subtype 1'. (B) Baseline anxiety-impulsivity relationship in subtype 2'. (C) Longitudinal associations between anxiety and lack of perseverance in subtype 1'. (D) Longitudinal associations between anxiety and sensation seeking in subtype 2'. Anxiety was measured by CBCL-Anxiety Problem. Impulsivity was measured by sub-facets of UPPS-P. Threshold of significant p-value was 0.05. nu, negative urgency; pu, positive urgency; lope, lack of perseverance; lopl, lack of planning; ss, sensation seeking; anx, anxiety.
